# Estimating and forecasting the burden and spread of SARS-CoV2 first wave in Colombia

**DOI:** 10.1101/2021.01.15.21249818

**Authors:** Jaime E. Cascante Vega, Juan M. Cordovez, Mauricio Santos-Vega

## Abstract

Following the rapid dissemination of COVID-19 cases in Colombia in 2020, large-scale non-pharmaceutical interventions (NPIs) were implemented as national emergencies in most of the municipalities of the country starting by a lockdown on March 20th of 2020. Using combinations of meta-population models SEAIIRD (Susceptible-Exposed-Asymptomatic-Infected-Recovered-Diseased) which describes the disease dynamics in the different localities, with movement data that accounts for the number of commuters between units and statistical inference algorithms could be an effective approach to both nowcast and forecast the number of cases and deaths in the country. Here we used an iterated filtering (IF) framework to fit the parameters of our model to the reported data across municipalities from march to late October in locations with more than 50 reported deaths and cases historically. Since the model is high dimensional (6 state variable by municipality) inference on those parameters is highly non-trivial, so we used an Ensemble-Adjustment-Kalman-Filter (EAKF) to estimate time variable system states and parameters. Our results show that the model is capable of capturing the evolution of the outbreak in the country and providing estimates of the epidemiological parameters in time. These estimates could become the base for planning future interventions as well as evaluate the impact of NPIs on the effective reproductive number (ℛ_*eff*_) and the key epidemiological parameters, such as the contact rate or the reporting rate. Our approach demonstrates that real-time, publicly available ensemble forecasts can provide robust short-term predictions of reported COVID-19 deaths in Colombia. This model has the potential to be used as a forecasting and prediction tool to evaluate disease dynamics and to develop a real time surveillance system for management and control.

## 1 Introduction

Coronavirus disease 2019 (COVID-19) pandemic emerged in December 2019 caused by the virus SARS-CoV-2 [1, 2]. This pandemic started in Wuhan-China, but it quickly spread to several countries worldwide [1]. This rapid global spread of SARS-CoV-2 has caused an urgent need of readily-available forecasts of the spatio-temporal transmission patterns to inform risk assessment and planning instances. In Colombia, the first positive case for the novel corona-virus (SARS-CoV-2), was reported on March 6 of 2020 in Bogota D.C. Then, the virus has spread rapidly to several municipalities in the country, and as of October 29 21 of 2020, 71 municipalities reported historically more than 50 deaths. On March 20 the government declared a nation-wide lockdown to prevent the spread of the virus throughout the country. After the first lock downs, several non-pharmaceutical interventions including case isolating, contact tracing, quarantine of exposed persons, social distancing, travel restrictions, school, churches and workplace closures were in place in Colombia to reduce transmission of the virus [3, 1]. Although, some of these measures are still in place; the intensity of these restrictions have changed over time as a consequence of reopening attempts, generating changes in mobility and activity patterns. Thus, assessing the temporal variation of transmission in real-time for different regions of the country, based on human mobility, becomes essential for evaluating the possible effects of reopening the economy of the country. Nowcasting and forecasting COVID19 dynamic can also be used to do early detection of possible time periods or scenarios with high transmission intensity, and ultimately help the public health system to assess, intervene and formulate long term public health policies.

The ability to generate nowcasts, forecasts, and the early identification of epidemic events can help to develop early-warning systems, which is important to timely implement effective control policies in developing countries [4, 5, 6]. In recent years, the interest in generating real-time epidemic forecasts to help the control and management of infectious diseases has grown, prompted by a succession of global and regional outbreaks of infectious diseases such as Zika, Ebola among others [7, 8, 9]. The current availability of epidemiological and digital data streams, enhanced by process-based models that incorporate climate, demography, and mobility among other factors, can provide a basis to evaluate the impact and effectiveness of intervention strategies in changing environments [10, 11, 12]. Different studies have proposed a variety of mechanistic and statistical approaches to forecast seasonal and pandemic diseases [13, 14]. The limitation of statistical models is that these approaches focus on associations and correlations in the epidemiological time series data, but it is not possible to disentangle the mechanisms behind disease transmission dynamics. [15, 13]. Opposed to mechanistic models which are based on biological mechanisms that underlies disease transmission, although forecasting with this models is often computationally intractable [14, 16, 10, 17, 18].

Recently, mathematical models and forecasting algorithms have been used to forecast and survey diseases such as Ebola [5], influenza [9, 10] and dengue [6]. These forecasting approaches combine assimilation techniques with dynamical transmission models, which allows to re-estimate parameters in real time; therefore their outputs would allow a rapid assessment and decision-making. Forecasting of infectious diseases is valuable tool that allows to understand disease transmission dynamics and to plan future interventions, although it can have biases due to assumptions in the short term value of key epidemiological parameters that mean to map real world setting or human behavior [19, 11]. In this paper, we used a population SEAIIRD model and a iterated filter algorithm to estimate and model the transmission of COVID-19 for municipalities that by the first week of October have reported more than 50 historical deaths. Parameter estimates in this paper are an important contribution for the understanding of SARS-CoV2 space-time dynamics, and combined these simulations with data assimilation techniques we are able to generate weekly real time now-casting and forecasting of this critical infectious disease in Colombia.

## 2 Methods

### 2.1 Data Description

We used daily reported cases (new infections) by the Instituto Nacional de Salud (INS) in Colombia, (this data is available in their Coronavirus web page [3]). Each new infection is identified by a unique case ID and has an associated notification date by the surveillance system (SIVIGILA), the symptoms onset date (reported by the patient to the health care provider) and diagnosis date (reported by laboratory after test confirmation). The epidemiological data also includes recovery date (when an infected patient tested negative for the virus), date of death, age, sex, municipality (county), department, type (imported from other country versus associated i.e., locally-acquired), location, if the patient is currently at home, hospital or ICU and the state/level of the disease (mild, medium or severe symptoms). The number of daily infected people (This number can be denoted by exposed, pre-symptomatic, symptomatic, or asymptomatic disease stage) depends on the testing effort, namely, the percentage of positive tests among all tests conducted on a given day. However, we assumed that most reported infections are from symptomatic individuals with severe or mild symptoms.

We also consider movement between municipalities Figure S2S2, for this we used movement data from Facebook into our models. Facebook ‘s Data for Good team developed and provides access to the Geoinsights portal to provide movement and population level data in response to crises. We used Facebook ‘s regular movement data, which aggregates the number of trips Facebook users make between locations over time [20].

### 2.2 Model description

We considered movement between municipalities as the movement in the meta-population. Our model is formulated as a discrete Markov process across days, and it assumes that susceptible individuals get infected at rate *λ*_*i*_ which we refer from now as the force of infection (**FOI**). In the model the **FOI** is proportional to the number of contacts between susceptible individuals (*S*_*i*_) and infectious individuals *I*_*i*_ at rate *β*_*t*_ and with asymptomatic individuals *A*_*i*_ at rate *σ*_*t*_*β*_*t*_. The model subdivides infectious stages in 3 classes: **i)** Exposed (E): Individuals who have the virus but cannot infect others. **ii)** Asymptomatic or individuals not reported as infected (A): Individuals who have the virus but do not present with symptoms or individuals who do not get tested for SARS-CoV2. **iii)** Infected: Individuals who are infectious for an average time of days before acquiring immunity. Moreover, we suppose asymptomatic individuals (with none or mild symptoms) are a proportion *α*_*t*_ of the total infections.

Our transmission model, assumes multiple locations are connected by human mobility, then populations are subdivided in *S*_*i*_(*t*), *E*_*i*_(*t*), *A*_*i*_(*t*), *I*_*i*_(*t*) and *R*_*i*_(*t*), which correspond to the susceptible, exposed, asymptomatic, infected, and recovered populations of a municipality *i* at time *t*. Importantly, we define patients with severe enough symptoms to be confirmed as documented infected individuals; while other infected people are defined as undocumented or asymptomatic infected individuals. We provide a parameter *β*(*t*) such as the municipal contact rate for transmission due to documented infected individuals. We assume that the transmission rate due to undocumented persons is reduced by a factor *σ* _*i*_ for asymptomatic individuals. In addition, *α* is the report fraction or the proportion of *I*_*i*_(*t*) individuals. *T*_*e*_ is the incubation time in days (time from infection to symptom onset for symptomatic individuals), *T*_*r*_ is the infectious period (time from symptom onset for symptomatic individuals or since exposure for asymptomatic individuals to recovery) also in days and *T*_*d*_ is the death period (time since exposure to death for individuals that are eventually gonna die). *M*_*ij*_(*t*) represents the human movement from municipality *i* to *j* at time *t* and is the amount of individuals in that time. Figure 2 shows the model diagram for the population dynamics of COVID-19.

In the meta-population model, the information on commuters is encoded in the size of sub-population *N*_*i*_. Specifically, we use *M*_*ij*_ to represent the number of commuters who live in location *i* and commute to location *j*. The transmission dynamics at each location follow the SEAIIRD model. To complete the transmission equations, we need to derive the population exchange across sub-populations caused by diffusive movement. The transmission equations follow the structure below: The transmission model equations follow the following structure, we use the 2020 DANE projection for the total population of the *i* − *th* municipality *N*_*i*_ [21].

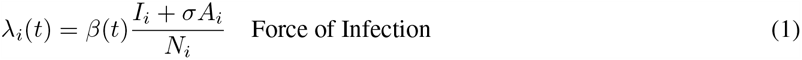

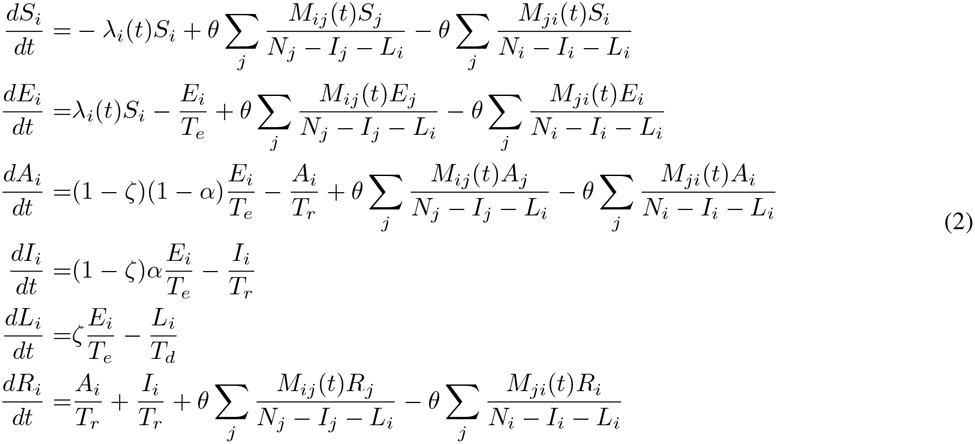

### 2.3 Parameter Estimation and forecasting

#### 2.3.1 Parameter inference

We parameterized the transmission model using the municipality-level incidence and death data reported from March 06 to October 11 2020. To estimate model parameters we use an Ensemble Adjustment Kalman Filter (EAKF), which is applicable to high dimensional models [22, 16, 10]. A iterative filtering approach to infer model parameters was used [23], this iterated filtering (IF)-EAKF framework has been used to infer parameters in large-scale models as network metapopulation models for other pathogens [23, 10, 16, 24]. This method starts by sampling different parameter combinations from a uniform distribution using latin-hypercube sampling with the prior ranges defined in Table S1. For addressing the surveillance system report issue we choose the prior fraction of reported cases *α* to cover almost all its domain. A similar range was used for the relative asymptomatic transmission for unreported individuals that have been shown to be mostly asymptomatic *σ* [16, 19, 25]. Importantly, in this case we assume that viral load in this sub-population cannot be greater than the viral load reported individuals *I*_*i*_ as has been assumed and estimated [1, 25].

Our initial conditions reflect the beginning of the lock-down in Colombia (March 20th). Here, we assume that infected individuals were not only imported to one principal administrative unit like Bogotá D.C. but to the principal Colombian districts and some surrounding municipalities. Hence we also capture the chance that the outbreak might be initiated in multiple municipalities due to community transmission. We assume equal initial conditions for all municipalities regardless of the number of cases reported in the first days of SARS-CoV detection. We assume that on the first day an infected individual was reported, there were 3 exposed individuals *I_i_*(0) = 1, *E_i_*(0) = 3 at time *T*_*i*_(0) this corresponds to the first day of reported cases by diagnosis date in each Colombian municipalities.

#### 2.3.2 Reporting Delay

Our transmission model does not explicitly account for the time lag between the infection and their notification. However, we consider a delay between the report of the symptom onset and the laboratory test result. To account for this notification delay, we mapped the simulated documented infections to the confirmed cases using a separate observation delay model. To estimate this lag period, *t*_*d*_, we examined the data and the distribution of the time interval to the event (in days) from the onset of symptoms to confirmation is well adjusted by a Gamma distribution (Figure S1 S1). In practice, during the simulation of the transmission model, for each new documented infection that goes from *E*_*i*_ to *I*_*i*_, a random number for *t*_*d*_ with a Gamma distribution is generated. In other words, this case is “reported” as a confirmed infection *t*_*d*_ days after the transition from *E*_*i*_ to *I*_*i,t*_. Therefore, the number of cases reported in one day accumulate as the model is integrated over time.

### 2.4 Real time nowcasting and forecasting

We used a data assimilation algorithm (Ensemble Adjustment Kalman Filter (EAKF)) to generate real-time forecasts for the epidemic. Kalman filters usually assume a Gaussian distribution for both the prior and likelihood. Therefore, the distribution of the system state can be fully parameterized by the first two moments (the ensemble mean and covariance) [10]. Based on this assumption, the posterior mean and co-variance is calculated through the convolution of two Gaussian distributions. However, by generating the prior using a non-linear model (e.g. the SEIRS model here), the model-filter system is able to accommodate the nonlinear system dynamics despite the linear assumption of the Kalman filter algorithm.

For the EAKF, the ensembles are updated deterministically, then the ensemble mean and covariance match their theoretical values exactly. Then, the higher moments of the prior distribution are preserved in the posterior. The EAKF also adjusts the unobserved state variables and parameters based on their covariance with the observed state variables. EAKF is a very suitable technique in problems like this on because its implementation is independent of the dynamical model. This allows both to stimulate and make short term predictions assuming particular scenarios which maps to parameters space [23]. Here we used multiple observations from different locations are used to optimize the model, by iterating over all observations sequentially and adjusting the entire state vector.

### 2.5 Metrics

To assess model performance we used different metrics to evaluate how the nowcasts and forecasts generated perform every week. First, we assessed the performance of our nowcasts at a weekly horizon. For example we use the posterior estimates prior to time *T* and forecast assuming time variable parameters were constant in the forecasting horizon and equal to me average estimated for the previous week *t* ∈ [*T* − 10, *T*] this forecast reasoning have also seen in [26] for more details in the forecast methods see section S.5 in Supplementary Material. Then we evaluated the forecast performance using different scores to measure the fit to the observations [27, 28, 29]. We implemented a probabilistic assessment of our forecasts calculating the sharpness of predictive distributions subject to calibration [29, 30]. Finally, to evaluate the performance of the models, the data were split up into two subsets, with each subset used for testing once while the other subset was used for training the models. Results of different performance measures for the model model evaluated weekly are depicted in table 3. By analyzing the results in Table 1 reported monthly, it is clear that the model forecast accurately predicts diseases dynamics one week in advance. Over July our model consistently was able to predict the number of cases in each of the municipalities, although there is much higher variance week-to-week in these forecast compared to national forecasts each week.

**Table 1:**
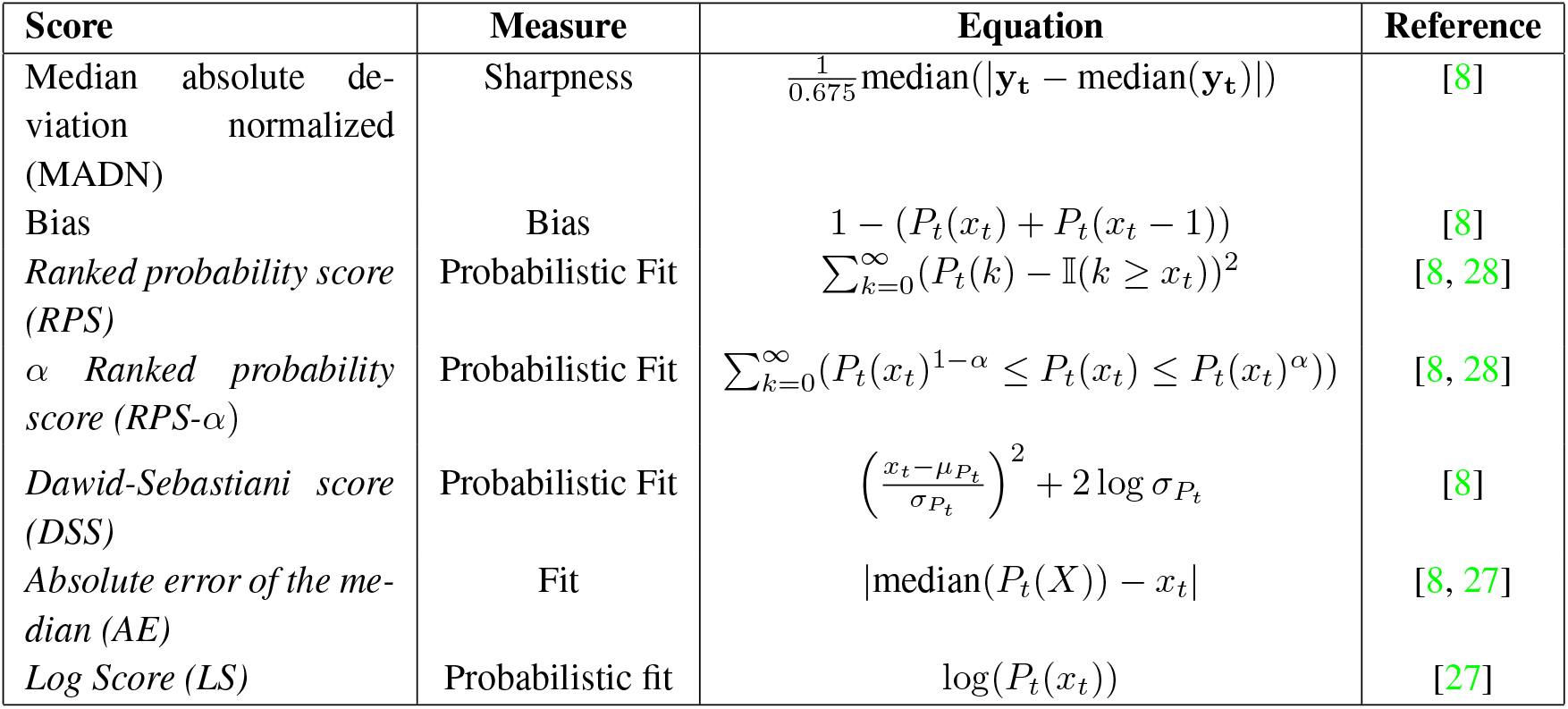
Summary of metrics used for evaluating the quality of both nowcast and forecast and their performance.

**Table 3:** Scores for evaluating probabilistic forecast. Scores are aggregated over the top-10 locations.

| Month | MADN | Bias | RPS | DSS | LS |
| --- | --- | --- | --- | --- | --- |
| May | 0.109 | 0.76 | 52.19 | 11.88 | -6.86 |
| June | 0.213 | 0.77 | 30.31 | 12.02 | -6.92 |
| July | 0.344 | 0.76 | 14.60 | 11.96 | -6.90 |
| Aug | 0.494 | 0.72 | 17.57 | 11.88 | -6.86 |
| Sep | 0.695 | 0.66 | 55.39 | 11.85 | -6.85 |
| Oct | 0.98 | 0.63 | 104.29 | 12.09 | -6.96 |

**Sharpness** is the ability of the model to generate predictions within a narrow range of possible outcomes. It is a data-independent measure, so it is purely a feature of the forecasts themselves as shown in table 1. To evaluate sharpness at time *t*, we used the normalized median absolute deviation about the median (MADN) of the prediction at time *t* [30]. Here the model forecast performances were averaged across the weekly estimates and reported for each month. We also assessed the **bias**, in other words if the model systematically over or under-predict, we defined the forecast bias at time *t* as shown in Table 1 [30]. An unbiased model would have *B*_*t*_ = 0 whereas an biased model would have *B*_*t*_ *>* 1 if the model overestimate at time *t* and *B*_*t*_ *<* −1 if the model under predict at time *t*. We say the model systematically over predict if *B*_*t*_ *>* 1 averaging across the times series. Similarly the model under predict if *B*_*t*_ *<* −1 in average. Finally, we evaluate a ranked probability score (RPS), which reduces to the mean absolute error if the forecast is deterministic [8]. and the coverages (CP) probabilities at confidence intervals of 95% and 50%. This score considers the number of observation that fall inside the specified model envelope [27].

## 3 Results

Our model inference approach was applied to three different periods of the epidemic; first to the period before the strong lock-down (March 03 to March 20), then during lock down period (March 21th to May 1st 2020) and finally the period of relaxation and progressively reopening (May 2nd to Oct 11th 2020). Parameter estimates for these three periods are reported in Table 2. Our estimates for the infectious period and latency period are ∼ 2.66 consistent with the period estimated in the literature (95%*CI*) [16]. In addition, the transmission rate *β*, and the report rate *α* at 95%*CI* are consistent with values assumed [1] or in estimated values [31, 32, 16].

**Table 2:** Estimated parameters in three different moments of the epidemic. Before the country level imposition of strict mobility restrictions. During the strict mobility restrictions and after relaxing mobility restrictions.

|  |  |  | Before lock-down | During lock-down | Lock-down relaxation |
| --- | --- | --- | --- | --- | --- |
| Parameter | Description | Units | Mean (95% CIs)<br>06-March - 20-March | Mean (95% CIs)<br>16-April - 15-May | Mean (95% CIs)<br>15-May - 11-October |
| $\beta$ | Contact rate | Days<sup>-1</sup> | 0.95 (0.93,0.97) | 0.94 (0.91,0.98) | 0.94 (0.90,0.98) |
| $\sigma$ | Relative asymptomatic transmissibility | - | 0.30 (0.29,0.32) | 0.30 (0.27,0.32) | 0.28 (0.26,0.30) |
| $\alpha$ | Report fraction | - | 0.54 (0.51,0.55) | 0.54 (0.49,0.61) | 0.55 (0.52,0.59) |
| $\theta$ | Movement report | - | 1.38 (1.37,1.40) | 1.38 (1.36,1.41) | 1.39 (1.35,1.42) |
| $T_e$ | Incubation period | Days | 2.62 (2.56,2.68) | 2.62 (2.52,2.72) | 2.57 (2.43,2.70) |
| $T_r$ | Infectious period | Days | 2.41 (2.35,2.47) | 2.38 (2.28,2.47) | 2.36 (2.27,2.46) |
| $T_d$ | Infectious period | Days | 2.41 (2.35,2.47) | 2.38 (2.28,2.47) | 2.36 (2.27,2.46) |

Comparisons between model simulations and data are shown in Fig. 3 for the national level. This figure shows simulations of reported cases using the best-fitting model parameter estimates and their confidence intervals. These results from the stochastic simulations show that our model is able to capture the temporal dynamics of the epidemic. In addition, the best-fitting model captures the space-time pattern of infections with the novel coronavirus disease 2019 (COVID-19) in different municipalities in Colombia as shown in Figure 3 for the time pattern in Figure 1.

**Figure 1:**
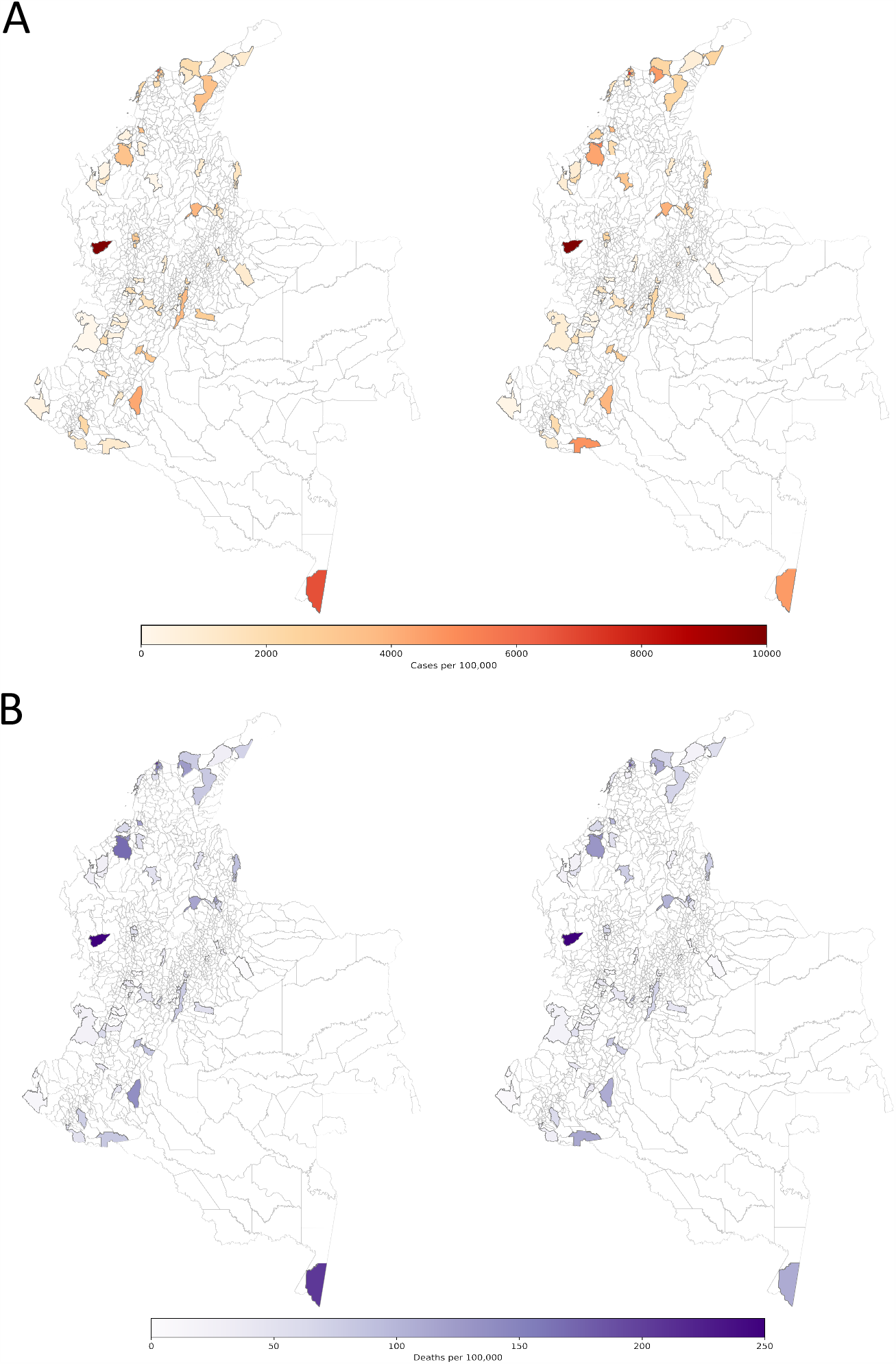
**(A LEFT)**: Cumulative observed cases of COVID19 by diagnosis date. **(A RIGHT)**: Cumulative estimated cases by the nowcasting in the EAKF metapopulation model. **(B LEFT)**: Cumulative observed deaths of COVID19. **(B RIGHT)**: Cumulative estimated deaths by the nowcasting in the EAKF metapopulation model.

**Figure 2:**
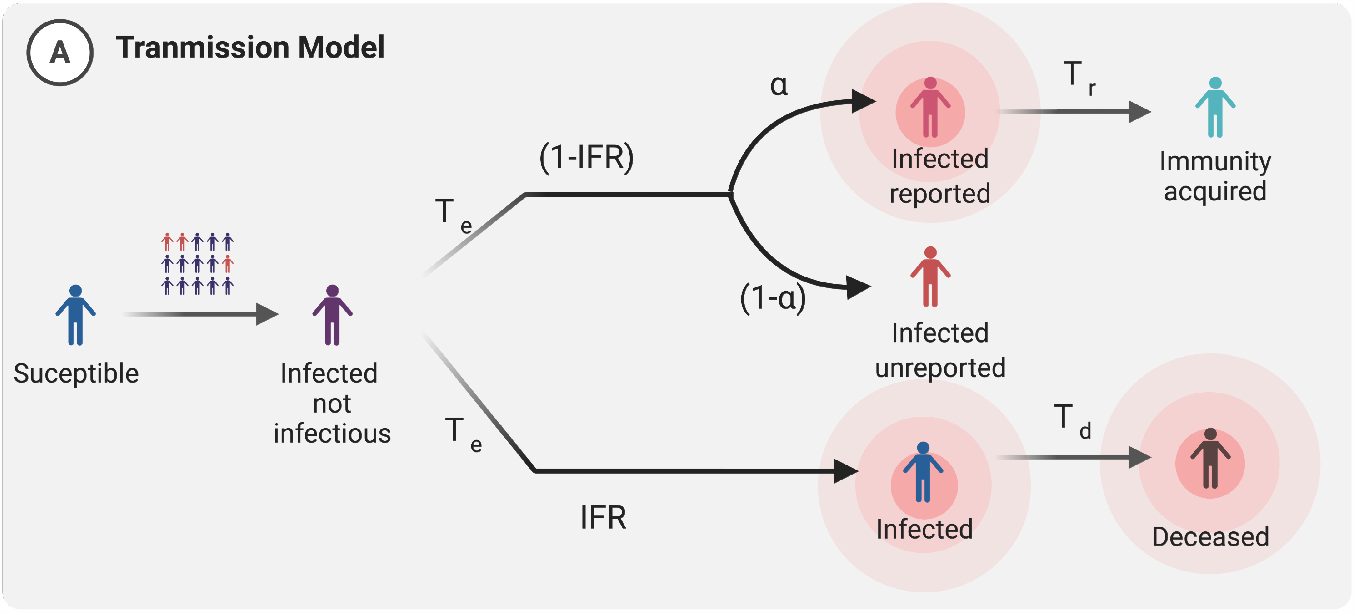
Meta-population SEAIIRD model. **A)** Schematic representation of the spatially explicit epidemiological model, where population is subdivided in Susceptible (*S*), Exposed (*E*), Asymptomatic or mild infections (*A*), Infected (*I*), Infected individuals that eventually are gonna die (*L*) and Recovered (*R*). This captures the local transmission dynamics in every municipality, importantly yellow compartments represent individuals who do not move within municipalities. **B)** Schematic of meta-population model, connections between municipalities.

**Figure 3:**
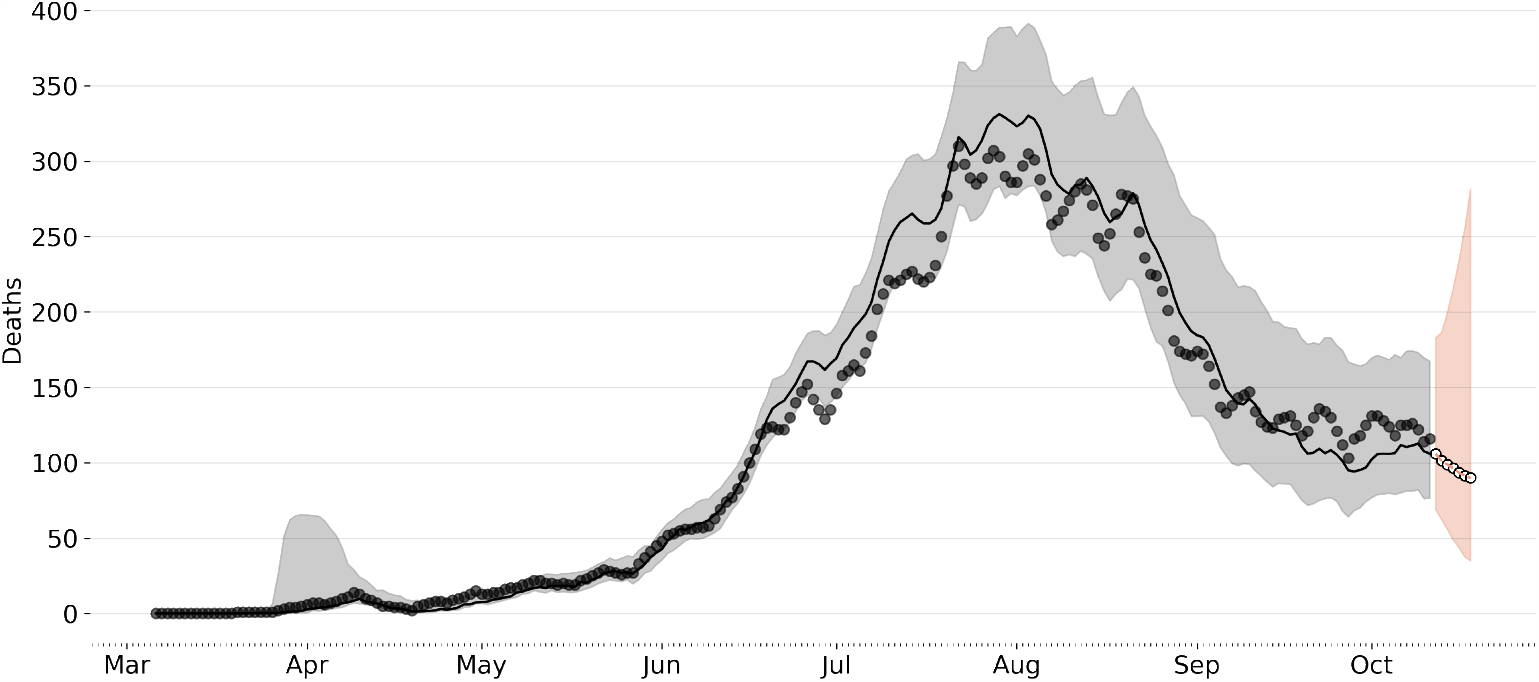
National Forecast (Aggregated municipal level forecast). This aggregation is the sum of all the deaths predicted for each municipality. Black line represents the median of the now-casting, the gray dark points are the daily deaths and the light gray envelope represents the 95% confidence interval. The orange white-dotted line represents the forecast assuming the parameters as the mean of the last week. Again the light orange envelope represents 95%.

Our median estimate of the effective reproductive number (*R*_*eff*_) is presented in figure 4. This is equivalent to the basic reproductive number, *R*_0_, at the beginning of the epidemic was around 2.2 [95 % credible interval (CI): 2.21 − 2.32] which coincide with the reported ℛ_0_ reported for Colombia for COVID19 [**?**]. indicating that this number has been always above 1 for COVID-19 in the country, suggesting a high capacity for sustained transmission (Table 1 and Fig. 1D). Importantly, reductions in *R*_*eff*_ are associated with the lock-down measures during the April, with sustained increases in this number after the reopening. Figure 4 also show the value of the parameters which *R*_*t*_ depends on (*β*_*t*_, *σβ*_*t*_ and *α*). Noteworthy, variation in the contact rates (*β* = *β*_*t*_) closely match the trajectory of *R*_*eff*_ and the cases in the country. There is a decreasing trend in the number of detected infections, and important variations in the fraction of asymptomatic cases, which could be causing the most events of infection. We are able to compute the effective reproductive number *R*_*eff*_ as the case reproductive number *R*_*t*_ times the fraction of susceptible individuals in the population *S_t_/N* where 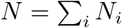 for every municipality, this can be seen in Figure S4 and S3.

**Figure 4:**
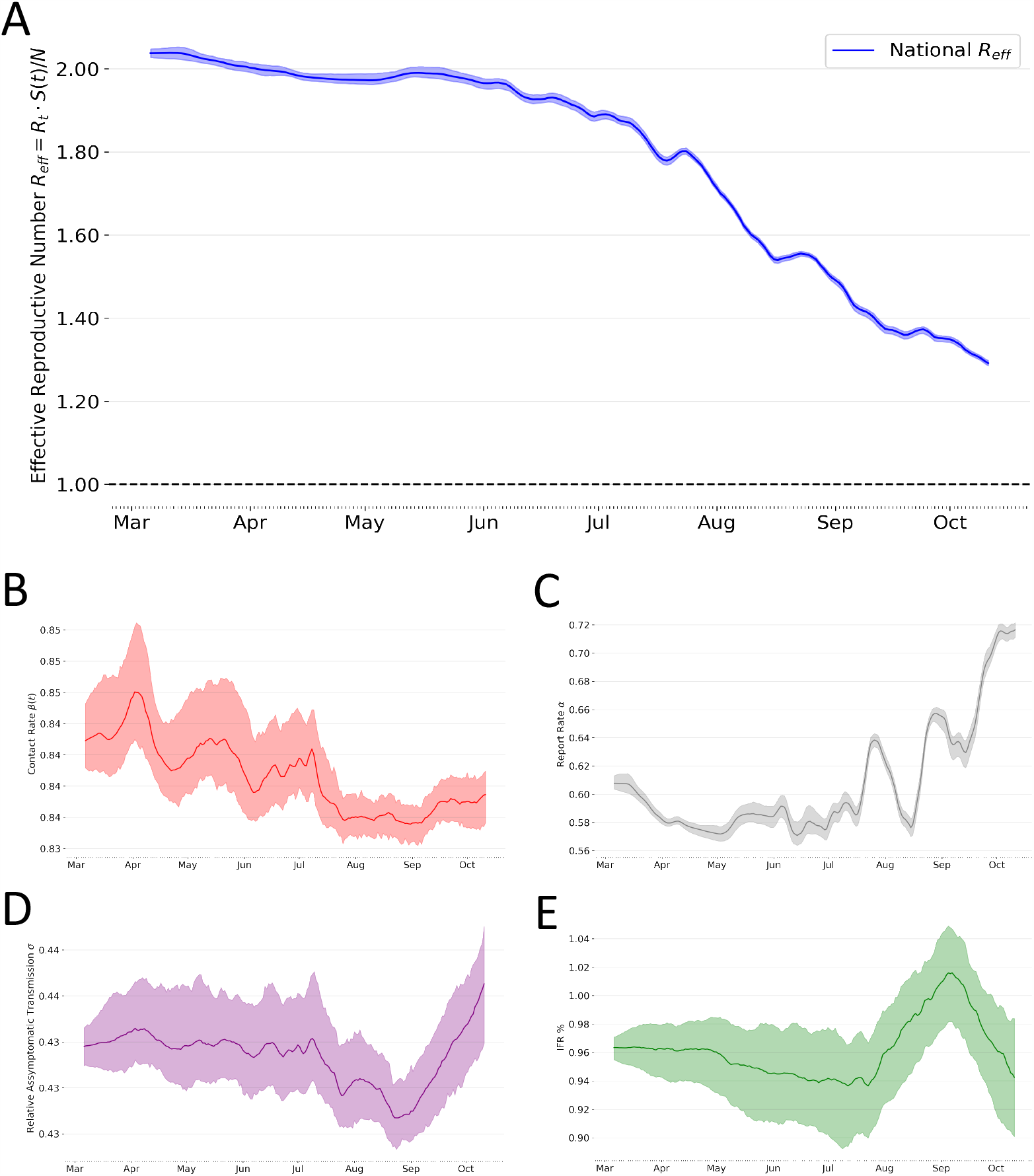
For all figures the lighter envelopes represent the 95% confidence interval and line represents the median estimate. **A:** National effective reproductive number computed as the mean of every municipality 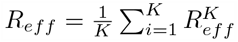; lighter envelope represent the the 95% confidence interval. **B:** Time variable contact rate *β*(*t*) lighter envelope represents the 95% confidence interval. **C:** National time variable report rate *α*. **D:** Relative Asymptomatic transmissibility *σ*. **E:** Infect Fatality Risk (IFR %) *ζ*

Figure 5 show forecastings for different representative regions of Colombia (the remaining regions are reported in the supplementary information). Our approach captured the temporal variation in the data at local scales, where most of the observed deaths in the plots fall within the confidence intervals generated with the model. The orange fringe shows weekly forecasting for the diseases.

**Figure 5:**
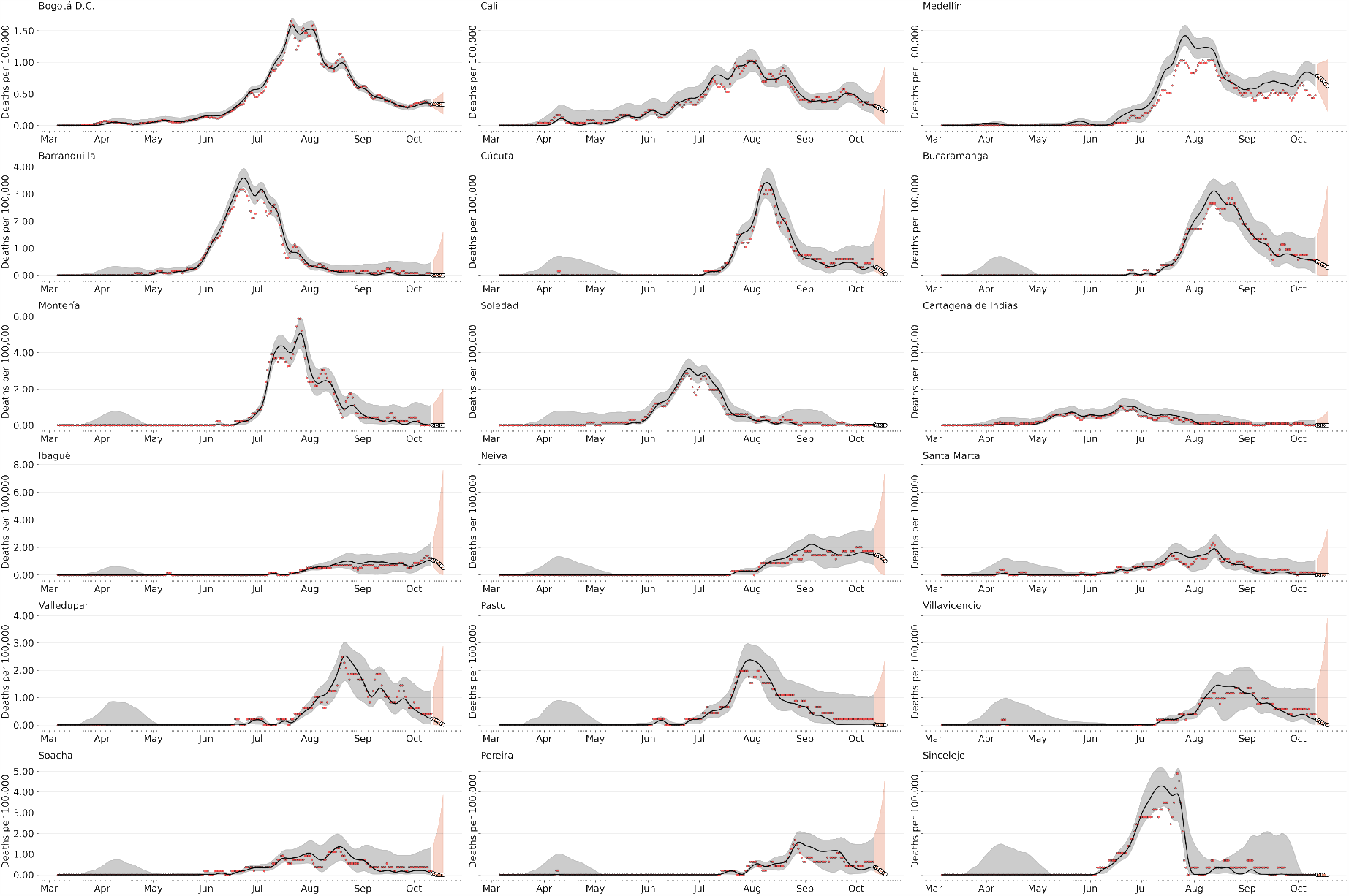
7 day death-forecast municipalities with more reported deaths by early October. Black line represents the median of the now-casting, the gray dark points are the daily deaths and the light gray envelope represents the 90% confidence interval. The orange white-dotted line represents the forecast assuming the parameters as the mean of the last week. Again the light orange envelope represents 90%.

## 4 Discussion

Our model generate forecasts that accurately predict the dynamics seen in the reported cases. Specifically, we are able to predict the daily cases and the epidemic dynamics one week in advance. Being able to forecast the daily cases and deaths allows for the proper prioritization and resource allocation by the public health authorities. The EAKF also allows parameter estimation, which is extremely useful to characterize the local transmission dynamics in the country and support the different modeling exercises to generate diseases trajectories in the long term. Standard epidemic SEAIR-type models implement a compartmental description under the assumption of homogeneous mixing of individuals [22]. Our results underscore the importance of short term forecasts in 1 week horizon, since the future of ongoing epidemics is so sensitive to parameter values that change over time, then predictions are only meaningful within a narrow time window, much as what we are used to in weather forecasts and recently in COVID19 deaths [33, 34]. We demonstrated that more detailed models using SIR-like with a meta-population structure incorporating mobility desegregate the effect of commuted usually incorporated in the contact rate *β* in the predictions and provide important insight about the dynamics. Also, considering a more realistic modeling approach where spatial heterogeneity due to time-varying disease onset times, regionally different contact rates, and the time-dependence of the contact rates is incorporated in the model provided important information in this exercise [35].

Our median estimates for the latency and infectious period, *T*_*e*_ and *T*_*r*_ respectively, are ∼ 2.66 and ∼ 2.4 days, and remains constant for most of the period fitted to the data. The parameter *α* asses the fraction of cases that is reported in the country, we found that just 55% of cases are reported. This estimate reveals a high rate of undocumented infections: 45% as have also been showed with different estimates around the globe [16]. Moreover the estimated time variable parameters (View Figure 4) generally agree with estimated parameters in the literature.

This assumption might not hold, as it seems that pre-symptomatic individuals might have higher viral load than symptomatic ones hence infect more [36]. However different models assume asymptomatic individuals infect less than symptomatic ones [37, 1, 38, 12] and transmission model parameter estimation also suggest this [32, 16].

We have demonstrated the potential of sequential data assimilation for COVID-19 dynamics at regional level and in combination with stochastic epidemiological models. By using an ensemble adjustment Kalman filter [22], we successfully have determined the contact parameter from simulated data and obtained reliable estimates from empirical data. Importantly, a characterization of the heterogeneity in the transmission parameter (beta) is the most critical free parameter in the stochastic SEAIIRD model, since other parameters (mean exposed and infectious duration or incubation period) can be extracted from the literature given that are intrinsic parameters of the disease [16, 37]. Given that our transmission rate is estimated in time and this parameter is directly related to the basic reproductive number ℛ_0_ [35], our approach becomes a useful method for statistical inference of the effective reproductive number (*R*_*eff*_).

Our results are the first estimates for the country and the only estimation of the under-reported or asymptomatic infections. These findings provide a baseline in Colombia for an assessment of the fraction of undocumented infections and their relative infectiousness. These results describe the transmission dynamics in Latino America, one of the regions with the highest attack rates and possible where the pandemic has impacted the most. Interestingly, our approach allows us to evaluate the impact of changes in interventions, viral surveillance and testing on the reported fraction. Here we found that on average the 40% of infections went undocumented could, however, shift in this fraction show their dependency on the time of the peak, the surveillance effort and the spatial heterogeneity. Our findings also indicate that a increase in the identification and isolation of currently undocumented infections is needed to fully control SARSCoV-2. Recent evaluation of forecast in the United States have shown the importance of including both probabilistic and point estimates metrics to assess both forecast accuracy (distance to observed data) and quality (coverage of forecast distribution) [26]. Therefore evaluation of the forecasting performance with different score measures show that our epidemiological model and the inference method has the capability of accurately predicting the number of cases in the country one week in advance, as reported results by month in table 1.

There are aspects with respect to these fore-castings that must be noted and considered during interpretation. First, the model is optimized using observations of both cases by confirmation date and deaths through October 11, 2020; however, because of this long delay between infection acquisition and case confirmation, any flattening of the curve due to these effects is not yet apparent in observations nor communicated to the model during optimization. Second, the spam to which this model has been optimized is highly variable in space and time, due to differences in contact behavior, population density, control measures, introduction of first case and testing practices. These differences in space and time make the fitting of any model of this scale challenging. In this changing context, it is very important to reiterate that it will take 10-14 days before the effects of real-world interventions—any flattening of the curve weeks—become apparent. Hidden infections create, after recovery, an unknown reduction in the number of susceptible that slows down epidemic dynamics; such an effect is currently not included in our current model. However, it seems compatible with our framework to extend the SEAIR model by an additional class of undocumented infected individuals.

Our model have four important assumptions on the SARS-CoV2 spread: **i)** We directly assume the delay from the infection to the report date fitting a Gamma distribution as shown in Figure S1 in Supplementary Material. This assumption directly assesses the challenge of reconstructing the time series of new infections, as observations occur long after the moment of transmission. [35]. **ii)** The model and the parameter inference setting let us to estimate time-variable contact rates for both reported individuals and asymptomatic/mild infections, which directly account for the mobility restrictions imposed to reduce the transmission. **iii)** The model also assumes time-variable asymptomatic/mild infections fraction, which accounts for the possible high number of asymptomatic infections in the country [37, 16].

Our work demonstrates the importance of using mechanistic models biologically motivated, and therefore have parameters that relate to well-established theory and can be interpreted by experts in the field. Although, using mechanistic models allow us to understand aspects of the dynamics behind the temporal patterns, we recognize that statistical models may prove more effective at using past observed trends to forecast the future. Many statistical models were designed to be either more flexible or parsimoniously parametrized, meaning that they may be able to more easily capture dynamics common to infectious disease time-series such as auto-regressive correlation and seasonality. In the case of these emergent pandemic, where limited data is available, mechanistic models may be able to take advantage of assumptions about the underlying transmission process, enabling rudimentary forecasts even with minimal data. On the other hand, many statistical models without assuming a mechanistic structure rely on past data to be able to make forecasts. That said, any forecasts made in settings with limited data must be subjected to rigorous sensitivity analyses, as such forecasts will necessarily be heavily reliant on model assumptions. In summary, our approach becomes a very useful tool for the country to understand the dynamics and estimate effects with comparatively little data at the level of regions, not an entire country.

## Data Availability

https://www.ins.gov.co/Noticias/Paginas/Coronavirus.aspx

## 5 Acknowledgments

We are grateful to the Instituto Nacional de Salud for providing the covid19 data for the country. We also thank Dr. Sen Pei to provide some code and technical information in the early stages of the project. JMC and MSV were supported by IDRC-Uniandes Grant 109582-001. We Thank Alejandro Feged, Vladimir Corredor for discussion about the paper, and Pallavi Kache and Carlos Bravo for initial review of the draft in early stages.

## Supplementary Material

### S.1 Basic Reproductive Number

For computing the Basic Reproductive Number we use the Next Generation Method (NGM) which account for changes in states where individuals have the virus *E, A, I, L* we compute the basic reproductive number as follows, we define the transition rates F and removal rates *V*.

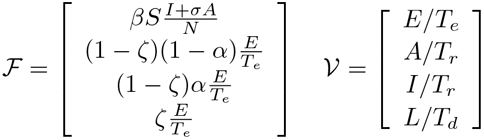

We then compute the transition and removal matrix F and V respectively as follows. And evaluate at the disease free equilibrium *S* = *N*.

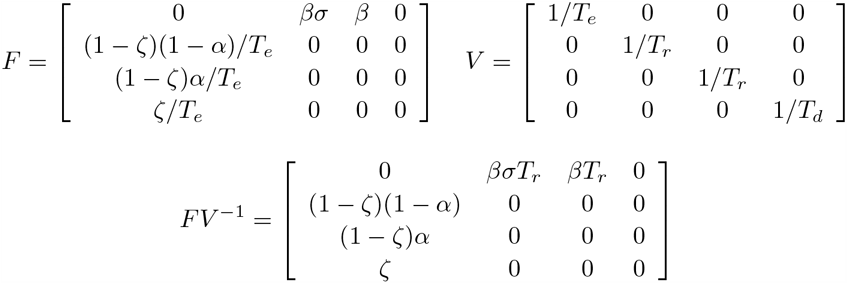

Then the basic reproductive number computed as the maximum eigen-value of *FV* ^*−*1^

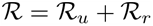

Where we define ℛ_*u*_ as the expected secondary infection due to under-reported individuals (recall that this we also believe this contribution is mostly due to asymptomatic infections which by definition are the individuals less captures by the surveillance system) and ℛ_*r*_ as the expected secondary infection due to reported individuals. This is consistent with the term involved in the proposed Force of Infection (FOI) in Equation 1, which only assume that unreported and reported individuals infect susceptible ones. Note that for the reproductive number do to under-reported individuals ℛ_*u*_ the fraction (1 − *ζ*) (1 − *α*) is the expected fraction of population entering the *A* compartment, here *zeta* is the infect fatality risk and *alpha* the fraction of reported individuals, where *σβ* is their transmission or contact rate and *T*_*r*_ is the average time expected in the *A* compartment. Similar for the reproductive number do to reported individuals ℛ_*r*_ we have (1 − *ζ*) *α* is the expected number of individuals entering the *I* compartment and *β* is their transmission rate and *T*_*r*_ is the expected time before acquiring immunity.

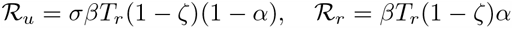

As we assume the contact rate is time variable *β* = *β*(*t*) we then compute the effective reproductive number in municipality *i* as, where *S*_*i*_(*t*) is the posterior sample of the estimated value of susceptible individuals at time *t* and *N*_*i*_ is the population in the municipalite *i*:

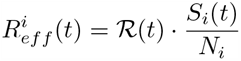

We then compute the national effective reproductive number as, for each one of the 71 municipalities that have reported more than 71 deaths by the date.

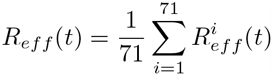

### S.2 Report Delay Distributions

We fit a Gamma distribution to the difference in days between symptom onset date and diagnosis date for addressing the report delay natural of the surveillance system using the SCIPY package available in Python [39]. The figure below shows some examples of the fitted Gamma distribution for the capitals of states/departments in Colombia. Note that using the fitted gamma distribution worth to model as probably by surveillance system or diagnosis laboratory reports an unusual number of cases is reported usually at 15 days. Therefore we recall the importance of used a fitted distribution rather than the empirical measured one. The importance of modeling this report delay have also been highlighted in current best practices for estimating the time varying effective reproductive number [40, 35].

**Figure S1:**
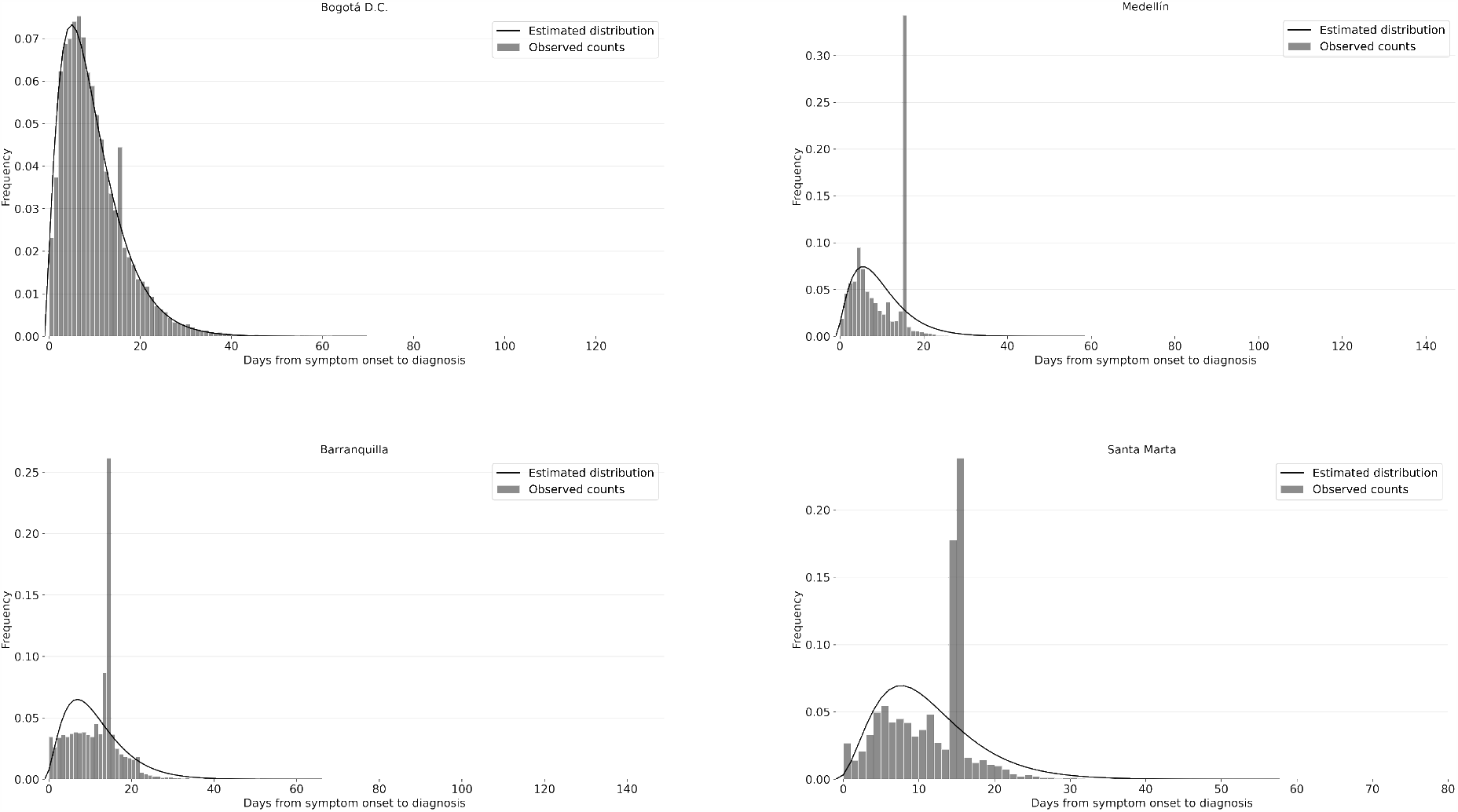
Fitted delay distributions for some municipalities. As seen is more frequent that the data is reported with a specific delay between symptom onset and diagnosis at 15 or 16 days (about 2 weeks) which we believe is due to return of diagnosis by the laboratories. Therefore we recall the importance of used a fitted distribution rather than the empirical measured one.

### S.3 Facebook Mobility Data

Commuters evolution from Facebook mobility data mobility in four different time periods.

**Figure S2:**
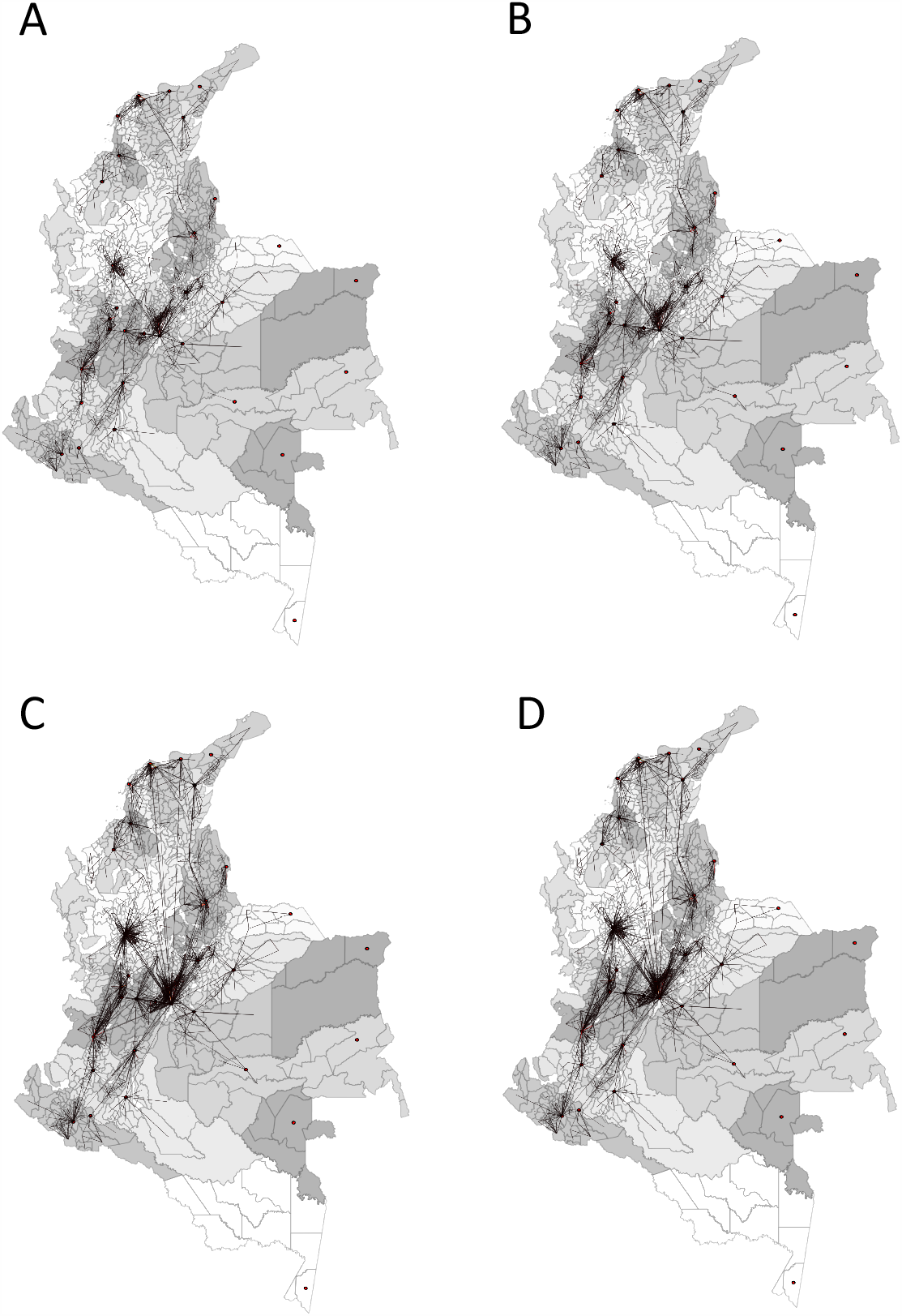
Facebook Mobility Data between municipalities, black lines represent commuters between a pair of municipalities and red dots are capitals of states/department of Colombia. States/Department are color-keyed as gray-scale in the map. Each map represent a specific time periods selected according to the national non-pharmaceutical interventions (NPIs) imposed in the country. **A** Average number of commuters in the strict national lockdown from 01-April-2020 (first available data) to 31-May-2020 first national reopening. **B** Average number of commuters from 01-June-2020 to 31-July-2020, two months after national re-opening. **C** Average number of commuters from 01-August-2020 to 31-October-2020, two month period four months after national re-opening. **D** Average number of commuters after 31-October-2020.

### S.4 Parameter Inference

For estimating the key local epidemiological parameters and therefore characterizing the disease spread of COVID19 we use flat uninformed priors as shown in table S1. For the infect fatality risk parameter (IFR) *3B6;* we search over all the IFR range reported for different countries and across ages [41, 42].

**Table S1:** Uninformative prior distribution on the estimated parameters for the epidemiological meta-population model

| Parameter | Prior Distribution | Reference |
| --- | --- | --- |
| $\beta$ | $\sim \mathcal{U}(0.75, 1.2)$ | - |
| $\alpha$ | $\sim \mathcal{U}(0.01, 1)$ | - |
| $T_e$ | $\sim \mathcal{U}(2, 5)$ | [43] |
| $T_r$ | $\sim \mathcal{U}(2, 5)$ | [16, 43] |
| $\theta$ | $\sim \mathcal{U}(0.5, 1.25)$ | [16] |
| $T_d$ | $\sim \mathcal{U}(7, 15)$ | - |
| $\zeta$ | $\sim \mathcal{U}(0.1\%, 2\%)$ | - |

### S.5 Operational Forecasting

For forecasting *t > T* where *T* is the last fitted day for all municipalities. We do not attempt to model or impose assumptions about future change in behavior that might impact the contact rate *β* or assume for example that testing is increasing resulting in increased report fraction *α*. We simply use the estimated values of the time varying parameters prior to the forecast horizon (we simple assume parameters will remain constant in the future). However for modeling temporal variation of behavior and based on the belief that behavioral changes occur smoothly in time we average over the last 10 days instead of using the last day estimates. We attempt to use more than one week as estimates might be impacted by report model of the surveillance system as has been showed in [40]. This approach have been similarly followed in [26]. We then assume for time variable parameters *θ*(*t*) that their value in the forecast horizon *t*_*f*_ follows Equation 3. Moreover for embedding this into the EAKF framework we treat each ensemble separately, therefore conserving parameter estimates within each ensemble member.

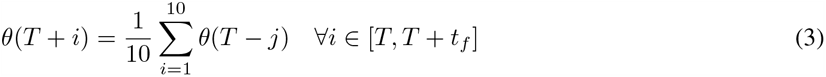

### S.6 Time estimates of the effective reproductive number

Median estimates of the effective reproductive number *R*_*eff*_ for the top 5 municipalities with more reported cumulative death by October.

**Figure S3:**
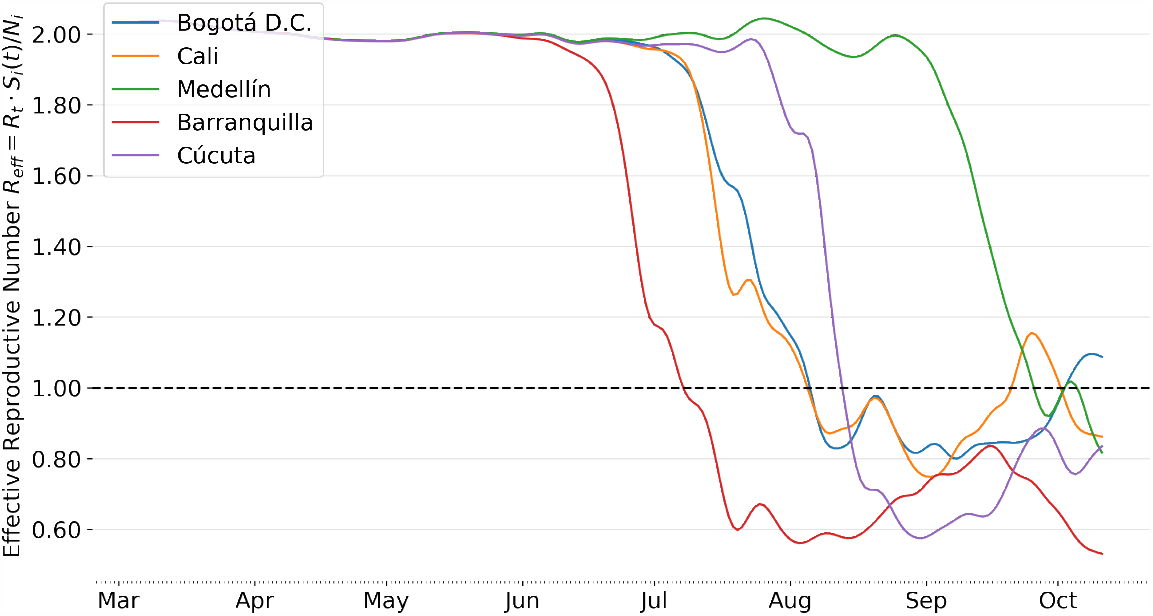
Median estimated of the effective reproductive number *R*_*eff*_ for the municipalities with more reported deaths by 11th October-2020.

### S.7 Spatial Distribution of the effective reproductive number

Spatial distribution of the effective reproductive number ℛ_*eff*_

**Figure S4:**
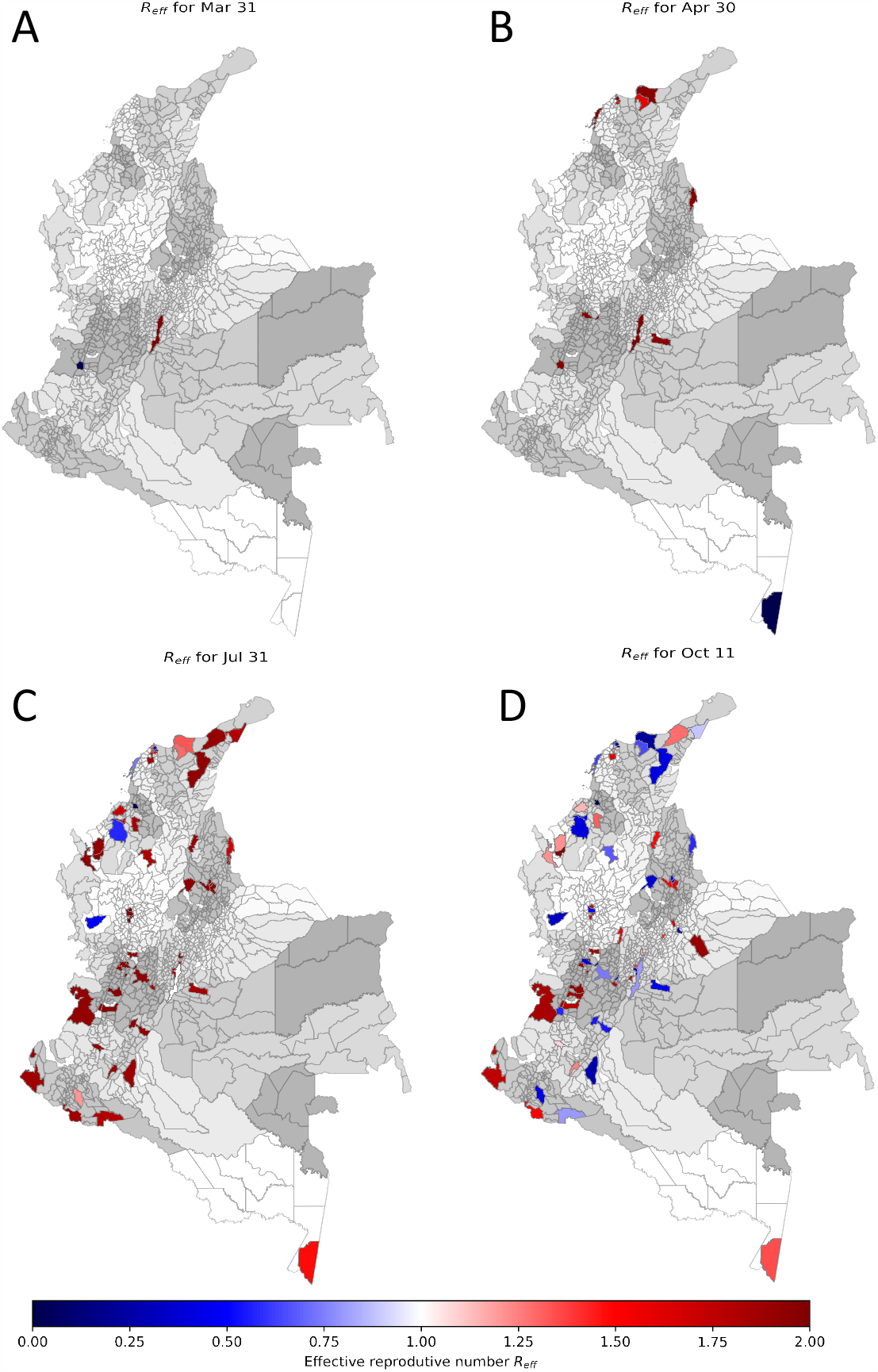
Effective Reproductive Number *R*_*eff*_ computed as time-variable reproductive number times the susceptible fraction estimated for each municipality 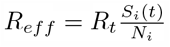. Color key: Red colors indicate median estimates *R*_*eff*_ *>* 1 while blue colors indicate *R*_*eff*_ *<* 1 magnitude is presented in colormap. **A)** *R*_*eff*_ for municipalities with reported deaths by March 31, one month since first reported case. **B)** *R*_*eff*_ for municipalities with reported deaths by Apr 31 2020, two months since first reported case. **C)** *R*_*eff*_ for municipalities with reported deaths by July 31 2020, five months since first reported case. **D)** *R*_*eff*_ for municipalities with reported deaths by October 11 2020, 8 months since first reported case.

